# Brain age as a surrogate marker for information processing speed in multiple sclerosis

**DOI:** 10.1101/2021.09.03.21262954

**Authors:** S Denissen, DA Engemann, A De Cock, L Costers, J Baijot, J Laton, IK Penner, M Grothe, M Kirsch, MB D’hooghe, M D’Haeseleer, D Dive, J De Mey, J Van Schependom, DM Sima, G Nagels

## Abstract

**Background:** Data from neuro-imaging techniques allow us to estimate a brain’s age. Brain age is easily interpretable as “how old the brain looks”, and could therefore be an attractive communication tool for brain health in clinical practice. This study aimed to investigate its clinical utility.

**Objectives:** To investigate the relationship between brain age and information processing speed in MS.

**Methods:** A ridge-regression model was trained to predict age from brain MRI volumetric features and sex in a healthy control dataset (HC_train, n=1690). This model was used to predict brain age in two test sets: HC_test (n=50) and MS_test (n=201). Brain-Predicted Age Difference (BPAD) was calculated as BPAD=brain age minus chronological age. Information processing speed was assessed with the Symbol Digit Modalities Test (SDMT).

**Results:** Brain age was significantly related to SDMT scores in the MS_test dataset (r=-0.44, p<.001), and contributed uniquely to variance in SDMT beyond chronological age, reflected by a significant correlation between BPAD and SDMT (r=-0.21, p=0.003) and a significant weight (−0.21, p=0.011) in a multivariate regression equation with age.

**Conclusions:** Brain age is a candidate biomarker for information processing speed in MS and an easy to grasp metric for brain health.

## Introduction

About half of the people with multiple sclerosis (MS) experience cognitive impairment(1), aggravating the impact of MS on their daily life and that of their caregivers(2). The most prominent of these cognitive difficulties is a slowing of information processing abilities(3), which literature suggests to be the key driver for deficits on other cognitive domains in MS(4). As difficulties with information processing speed (IPS) can induce problems such as falling, reduced quality of life and employment issues(4), and might negatively affect mental health in caregivers(5), timely identification is necessary. This requires regular and consistent follow-up in standard clinical care. Currently, neuropsychological testing remains the gold standard to detect cognitive problems(6), the most popular test in clinical practice for IPS being the Symbol Digit Modalities Test (SDMT)(7). Although the SDMT is a brief screening test(8), it is prone to practice effects(9).

An objective biomarker to diagnose IPS deficits might circumvent the aforementioned problem. Currently, predominantly structural brain characteristics, extracted from brain imaging techniques such as MRI, were found to be related to IPS(10). Yet more information can be extracted from an MRI than meets the eye. By using large datasets of brain images of healthy individuals, a machine learning model can be trained to estimate the age of a given brain. For a new brain image, the model will output the best guess of the age of that person’s brain, i.e. the “brain age”, which can look older or younger than the actual, chronological age of that person. The elegance of brain age is in its interpretable nature; it is easily graspable as how old a brain appears to be. In several brain disorders(11), among which MS(11–13), brains typically look older than those of their healthy peers.

On top of being an interpretable metric, recent evidence indicates that brain age could explain clinical symptomatology in MS. More specifically, increased brain age is associated to physical disability as quantified by the Expanded Disability Status Scale (EDSS)(11, 13) and nine hole peg test (9HPT)(12). Beyond physical disability, recent findings by Kaufmann et al 2019 in dementia suggest that brain age could explain cognitive disability as well, namely by finding that increased brain age was associated with lower scores on the Mini Mental State Examination (MMSE), independent of chronlological age(11).

In summary, brain age is an interpretable imaging-derived metric that is sensitive to MS-related pathology. However, it is currently unknown how brain aging is related to MS-specific dysfunction in processing speed. So far, efforts have mostly uncovered anatomical correlates by directly linking brain volumetry to information processing speed. Yet, people might be more easily capable of imagining “how old a brain looks” compared to “how voluminous a brain is”, posing an opportunity for a new communication tool in clinical practice that avoids medical jargon, which answers the desire of patients to be informed in plain language(14). In this study, we explore brain age as a tool for studying dysfunction in cognitive processing speed in MS on an international, multi-center dataset.

## Methods

### A. Data description

Data are described by subdividing them in HC_train, HC_test and MS_test, used to train and test the brain age decoding model as shown in figure 1. HC_train was constructed from a large sample of 1690 healthy control (HC) subjects from online open-source repositories. We refer to supplementary table S1 and figure S1 for a more detailed description. HC_test was collected as part of a study(15, 16) on understanding the neural origins of cognitive disturbances in MS, and for which both HC (HC_test, n = 50) and MS (n = 97) subjects were assessed. The MS subjects of this study were recruited at the National Multiple Sclerosis Center (NMSC) of Melsbroek, and were included in the MS_test dataset, which was further enriched with data from the Universitätsmedizin Greifswald (n = 104). For all data, T1-weighted Magnetic Resonance Imaging (MRI), sex and age at image acquisition were available. For the test datasets, Fluid Attenuated Inversion Recovery (FLAIR) MRI and results from the Symbol Digit Modalities Test (SDMT)(7) were available. The SDMT is a brief test, designed to measure information processing speed, that is attractive for its psychometric properties(17), quick administration(8) and its capability of predicting scores on other cognitive tests(18). Finally, MS_test data also contained EDSS, disease duration and type of MS. A summary of all data is available in table 1.

**Table 1.**
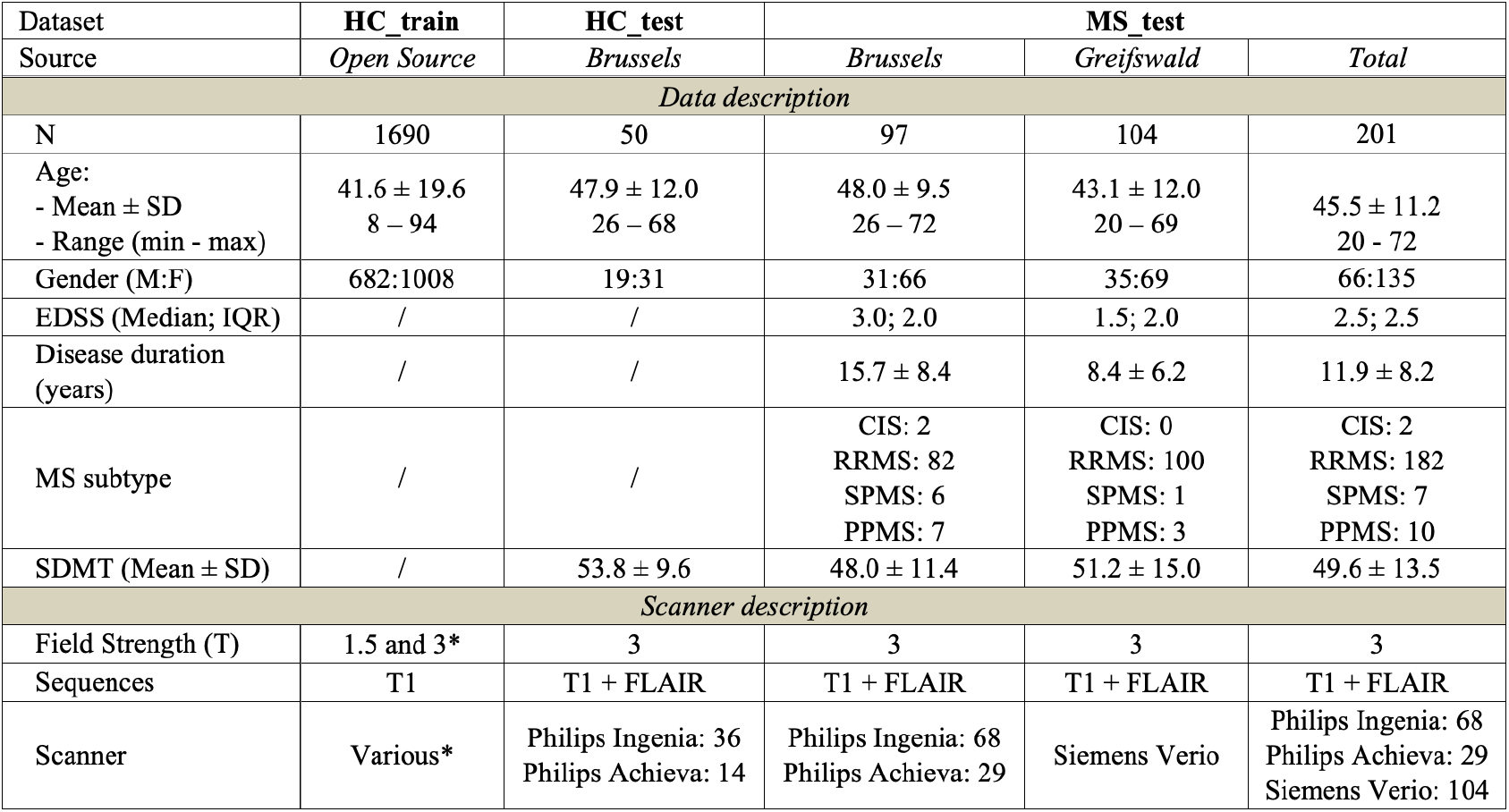
Data characteristics. Abbreviations: SD = standard deviation, M = Male, F = Female, IQR = interquartile range, T = Tesla, CIS = Clinically Isolated Syndrome, RRMS = Relapsing Remitting MS, SPMS = Secondary Progressive MS, PPMS = Primary Progressive MS. *We refer to supplementary table S1 for more details.

**Fig. 1.**
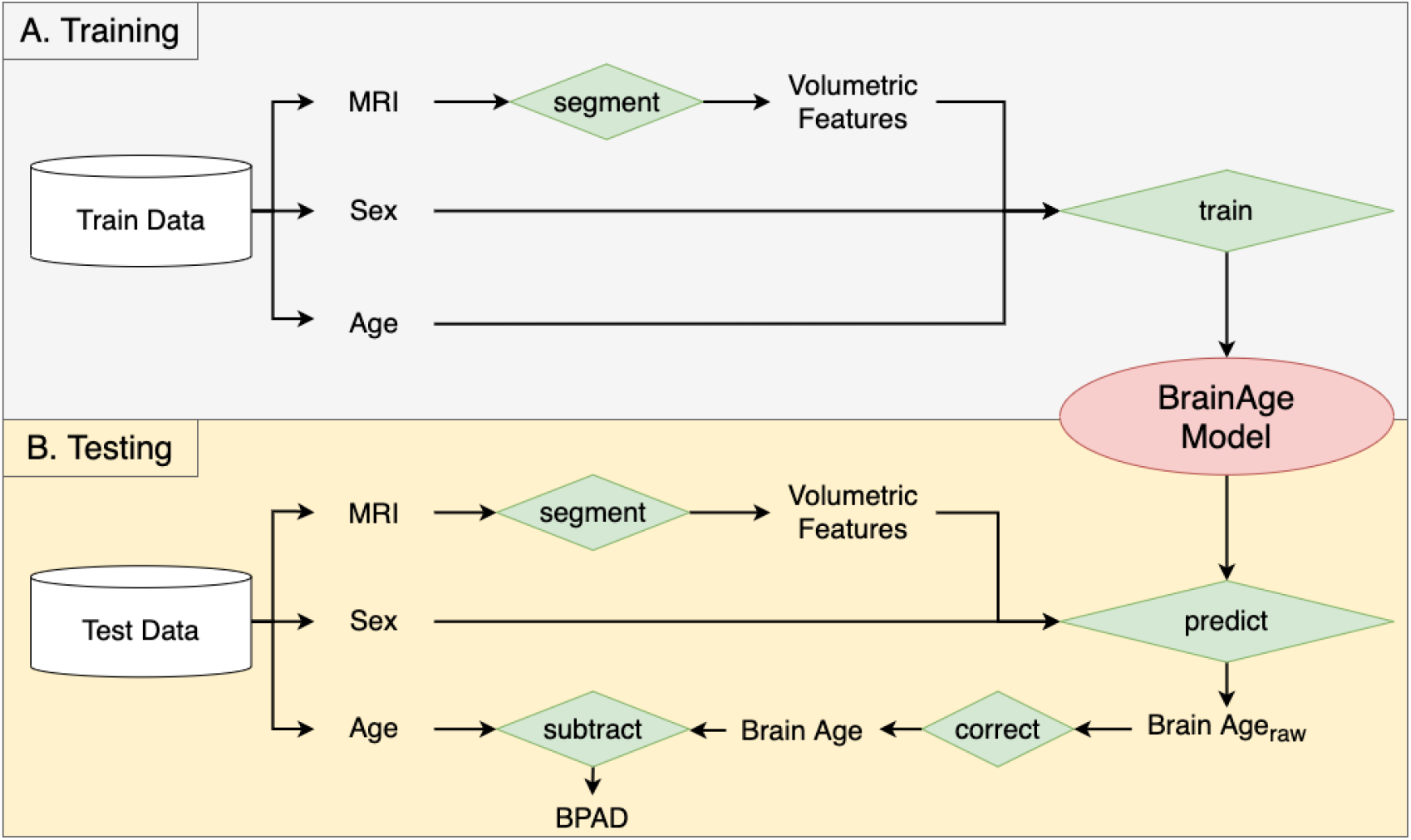
Brain age pipeline. The pipeline is subdivided in a Training phase (panel A) and a Testing phase (panel B), where “Train Data” refers to the HC_train data, and “Test Data” represents either the HC_test or the MS_test dataset. A silo-like shape represents a dataset, whereas green diamonds represent some kind of operation, specified by the text inside. Other text are either variables or images.

### B. Ethics

Participants of the BRUMEG study, Brussels, all provided their written informed consent prior to MRI assessment. The study protocol (B.U.N. 143201423263) was approved by the ethical committee of the UZ Brussel (Commissie Medische Ethiek (O.G. 016), Reflectiegroep Biomedische Ethiek) on 2015/02/25. For the patients from Greifswald, Germany, the study was approved by the ethics committee of the Medical Faculty of the University of Greifswald (BB159/18), and all participants gave their written informed consent. HC_train data consists of publicly available data originating from other projects, listed in supplementary table S1. Ethical approval was received by each project separately.

### C. MRI preprocessing and brain age pipeline

Several preparatory steps were involved in the construction of our brain age model. They are summarized below.

- *Brain MRI segmentation.* From the HC_train dataset, we extracted 3D T1-weighted MRI, sex and chronological age. Next, the T1-weighted MR images were evaluated by the FDA cleared ico**brain** software (version 4.4) of ico**metrix** NV (Leuven, Belgium). This is an end-to-end automatic software that segments and subsequently quantifies distinct regions of the brain. For a comprehensive description of the pipeline, originally published in Jain et al. 2015(19), we refer to Golan et al 2020(20). The pipeline relies on T1-weighted MR images, and when available, also uses FLAIR images to segment white matter lesions in the brain. These lesions are filled in the T1 image with white matter intensities, and the T1 image is subsequently segmented by fitting a probabilistic model for grey matter, white matter and cerebrospinal fluid image intensities. Ultimately, the pipeline generates volumes of the segmented regions, yielding a set of features, normalised for head size, that describe the brain’s morphology. These can be subdivided into general volumes (whole brain, grey matter, white matter, cortical grey matter, lateral ventricles), lobe-specific cortical grey matter (frontal, temporal, parietal, occipital) and subcortical volumes (e.g., hippocampus and thalamus (total, left and right)). Together with the subjects’ sex, this forms the total set of features used for the brain age pipeline. HC_test and MS_test were also segmented with ico**brain**, yielding the aforementioned set of features. The sole difference with HC_train is the additional availability of FLAIR images to perform lesion filling in the T1 image.
- *Ridge regression and selecting regularisation parameter lambda.* Along with chronological age, i.e. the target variable to be predicted, the total set of features (z-normalised) served as input for training a supervised machine learning model. Since the brain volumetric features in our data were mutually correlated, and ordinary linear regression could therefore result in large weights that are often indicative for overfitting(21), we used ridge regression. This method forces weights to shrink, a process that is controlled by regularisation parameter lambda. Lambda was established prior to training using a random search hyperparameter tuning.
- *Brain age correction.* According to Le et al. 2018, age-predicting models are prone to “regression towards the mean”, a phenomenon that results in overestimation of the brain age of younger subjects, and under-estimation of the brain age of older subjects(22). Inspired by Cole et al. 2020, we corrected for this bias by correcting brain age linearly for age(13). First, we estimated BrainAge_raw on HC_train by adopting 10-fold cross-validation. Second, we fitted a linear regression equation between the obtained raw brain ages and the respective chronological ages: *BrainAge*_*raw*_ = *β*_0_ + *β*_1_*ChronologicalAge* + *ϵ*. Herein, *β*_0_ and *β*_1_ represent the intercept and slope of the regression line, respectively, whereas the error term *ϵ* represents the residuals between data points and the regression line. *β*_0_ and *β*_1_ serve to correct each brain age predicted by our model using: *BrainAge* = (*BrainAge*_*raw*_ *− β*_0_)*/β*_1_. They were first used to correct the raw brain age estimates of HC_train.

Altogether, the preparatory steps yield the features and regularisation parameter lambda that are used for the training phase (figure 1, panel A) of the final brain age model, i.e. establishing the final feature weights on the full HC_train data using ridge regression. Next, we investigated whether the learned weights generalise to other datasets as well (figure 1, panel B). We first calculated raw brain age on the HC_test and MS_test dataset by calculating the weighted sum of the features (i.e. sex and brain volumes from ico**brain**) with their respective weight, which were then corrected by using the correction formula of preparatory step 3. In the remainder of this paper, “brain age” consistently refers to the corrected brain age.

### D. Statistical Analyses

Statistical analyses and visualizations were performed in Python and R. Significance level alpha was set to 0.05 for all reported test results. Where Pearson correlation was used, the distributions of the variables were checked for normality with a Shapiro-Wilk test. Raincloud plots were generated with use of the PtitPrince package(23).

First, to compare brain age, BPAD and chronological age distributions between MS_test and HC_test, we used a Mann-Whitney U test. To compare BPAD of both test sets with 0, we used a one-sample Wilcoxon signed rank test.

Second, we calculated the error of predicting age from brain images with the mean absolute error (MAE) between true and predicted age for every dataset. Next, to establish the association between brain age and SDMT in the MS_test data, we used Pearson correlation. To assess whether it contains unique information beyond chronological age, we used two approaches. First, we considered brain age and chronological age together in a multivariate linear regression equation: SDMT = *β*_0_ + *β*_1_*BrainAge* + *β*_2_*ChronologicalAge* + *ϵ*. Second, previous literature used the “Brain-Predicted Age Difference” (BPAD)(13). It quantifies brain age overestimation by subtracting chronological age from brain age. It was calculated in our test sets from the corrected brain age values (figure 1). Pearson correlation was used to establish the relationship between BPAD and SDMT in the MS_test data.

To test the complexity of the brain age measure, we compared it to the first principal component (*PC*_1_) of the feature space in the MS_test data by means of Pearson correlation.

## Results

### A. The brain age pipeline

Linear regression between brain age, obtained with 10-fold cross-validation on HC_train, and chronological age yielded the following correction weights: *β*_0_=8.32, *β*_1_=0.80. In the same dataset, the mean absolute error (MAE) between corrected brain age and chronological age was 7.84 years. The result of the brain age correction was added to the supplementary material as figure S2. In a next step, represented by panel A in figure 1, we fitted a final brain age model by using all available HC_train data for training. The model’s feature weights and lambda value can be consulted in table 2. To test its quality, we applied it to the HC_test and MS_test set, represented in figure 1 by panel B. The corrected brain age and BPAD values are summarised in table 3, and visually displayed for HC_test and MS_test in the raincloud plots of figure 2.

**Table 2.** The final brain age model’s characteristics. Both feature weights and the value of the regularisation parameter lambda are presented.

| Feature weights |  |
| --- | --- |
| whole brain | 2.035875 |
| grey matter | 2.892677 |
| white matter | -0.649640 |
| cortical grey matter | -13.212874 |
| lateral ventricles | 3.087088 |
| cortical grey matter – frontal lobe | -3.211897 |
| cortical grey matter – occipital lobe | 0.270182 |
| cortical grey matter – temporal lobe | -0.317308 |
| cortical grey matter – parietal lobe | -0.998446 |
| hippocampus | 1.237123 |
| hippocampus - left | -0.494472 |
| hippocampus - right | 0.775152 |
| thalamus | -1.641113 |
| thalamus - left | -0.950333 |
| thalamus - right | -2.208560 |
| sex | -3.528404 |
| Hyperparameters |  |
| lambda | 1.085003369566341e-05 |

**Table 3.** Outputs from the brain age pipeline. Corrected brain age and BPAD for the HC_train, HC_test and MS_test datasets.

|  | HC_train | HC_test | MS_test |  |  |
| --- | --- | --- | --- | --- | --- |
|  | *Open Source* | *Brussels* | *Brussels* | *Greifswald* | *Total* |
| N | 1690 | 50 | 97 | 104 | 201 |
| Brain Age (M ± SD) | 41.6 ± 22.0 | 46.8 ± 16.3 | 60.2 ± 16.8 | 62.4 ± 22.0 | 61.4 ± 19.6 |
| BPAD (Mean ± SD) | 0 ± 9.9 | -1.1 ± 9.4 | 12.2 ± 14.2 | 19.3 ± 15.0 | 15.9 ± 15.0 |

**Fig. 2.**
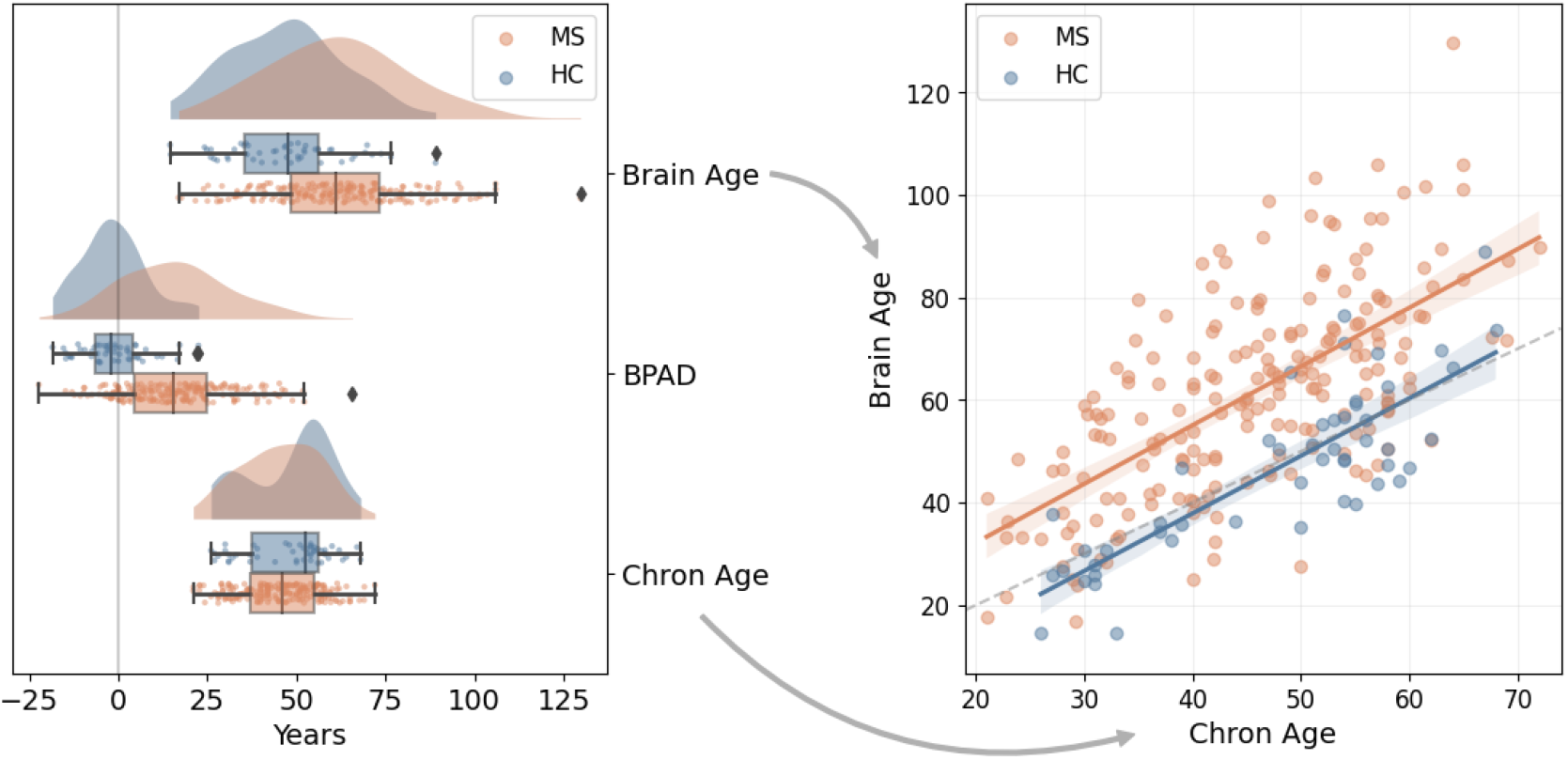
Group comparison between HC_test (blue) and MS_test (orange) for brain age, BPAD and chronological age. **Left:** the raincloud plots show the distribution of brain age, BPAD and chronological age for MS_test and HC_test. A reference line at x = 0 was included as visual aid. **Right:** the scatterplot shows the relationship between brain age and chronological age for MS_test and HC_test. The dotted line was added as reference, namely where brain age = chronological age.

1. Evaluation on HC_test data. The model predicted brain age with an MAE of 7.39 years. BPAD was not significantly different from zero (W = 509, p = 0.21), indicating that on average, brain age is similar to chronological age.
2. Evaluation on MS_test data. The model predicted brain age with an MAE of 17.5 years, and BPAD was significantly greater than zero (W = 1193, p < .001).

We additionally note that both brain age (U = 2870, p < .001) and BPAD (U = 1660, p < .001) were significantly higher in the MS_test data, compared to HC_test. Chronological age was comparable between both groups (U = 4336.5, p = 0.07).

### B. The relation between brain age and information processing speed

Brain age was significantly correlated to SDMT (figure 3, r = −0.44, p < .001). Moreover, brain age explained unique variance in SDMT beyond chronological age, which is reflected by a significant correlation between BPAD and SDMT (figure 4, r = −0.21, p = 0.003) and the significant weight (*β*_1_ = −0.21, p = 0.011) assigned to brain age when considering it in the multivariate regression equation *SDMT* = *β*_0_ + *β*_1_*BrainAge* + *β*_2_*ChronologicalAge* + *ϵ* (figure 5). Chronological age also contributed significantly to the model (*β*_2_ = −0.35, p < .001).

**Fig. 3.**
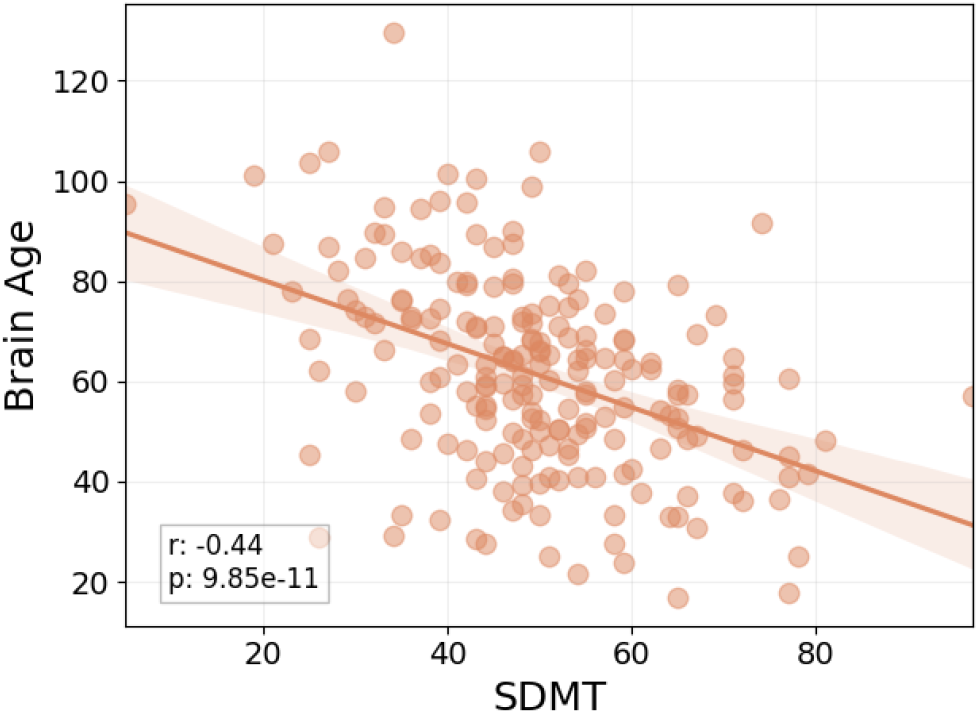
Scatterplot between Brain Age and SDMT in the MS_test dataset. The textbox describes the Pearson r statistic, along with the p-value.

**Fig. 4.**
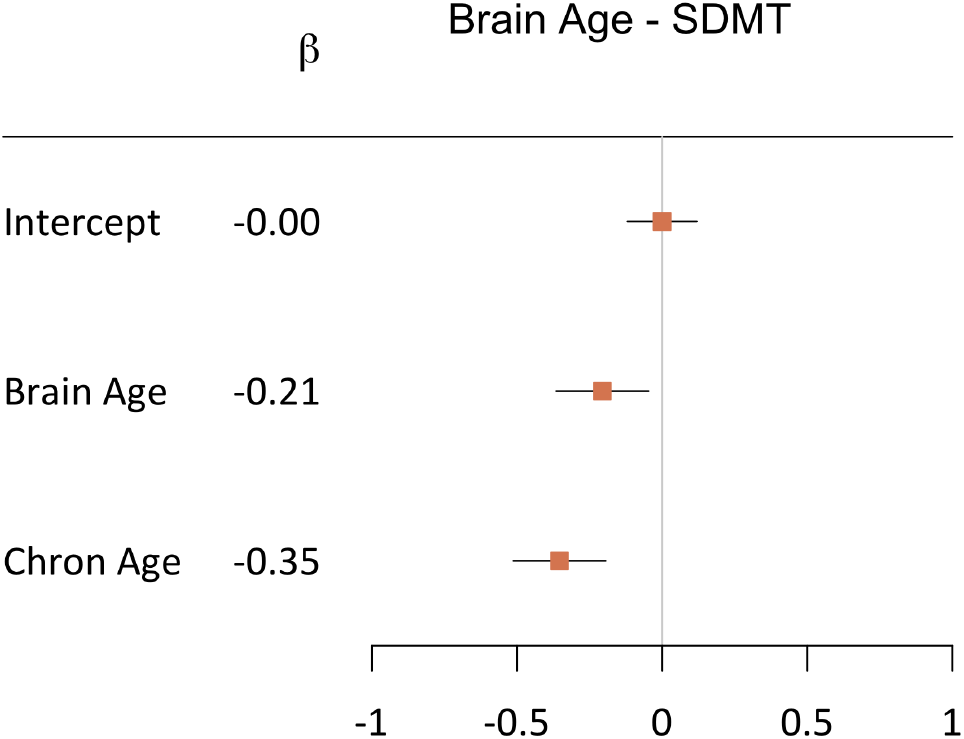
Forest plot visualising the significance of the weights (*β*_*n*_) in the linear regression equation *SDMT* = *β*_0_ + *β*_1_ *BrainAge* + *β*_2_ *ChronologicalAge* + *ϵ* in the MS_test dataset. The maximum likelihood estimates of the weights (*β*_*n*_) are represented by the orange squares, along with a 95% confidence interval (horizontal bar). If the latter does not include 0, the contribution of that feature to the model is considered significant. Brain age and chronological age contributed significantly (p = 0.011 and p < .001 respectively), whereas the intercept did not (p = 1.00).

**Fig. 5.**
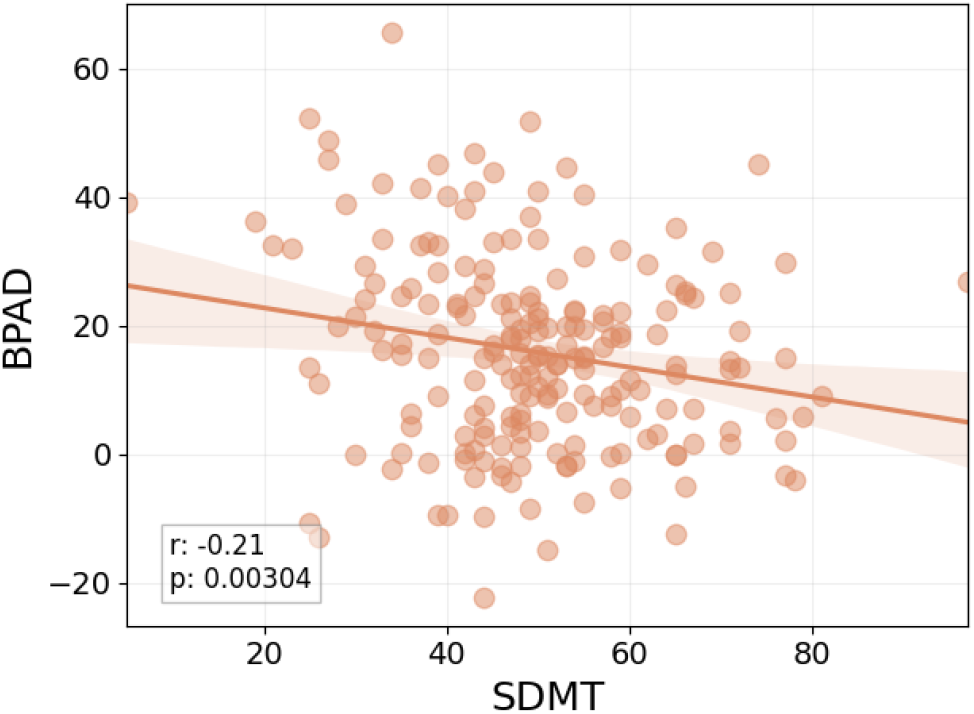
Scatterplot between BPAD and SDMT in the MS_test dataset. The textbox describes the Pearson r statistic, along with the p-value.

### C. The relation between brain age and brain volumetry

*PC*_1_ explained 85.3% of the total variance that is explained by the total set of features in the MS_test data. Figure 6 displays the relationship between *PC*_1_ and brain age, revealing a strong and significant linear relationship (r = 0.94, p < .001).

**Fig. 6.**
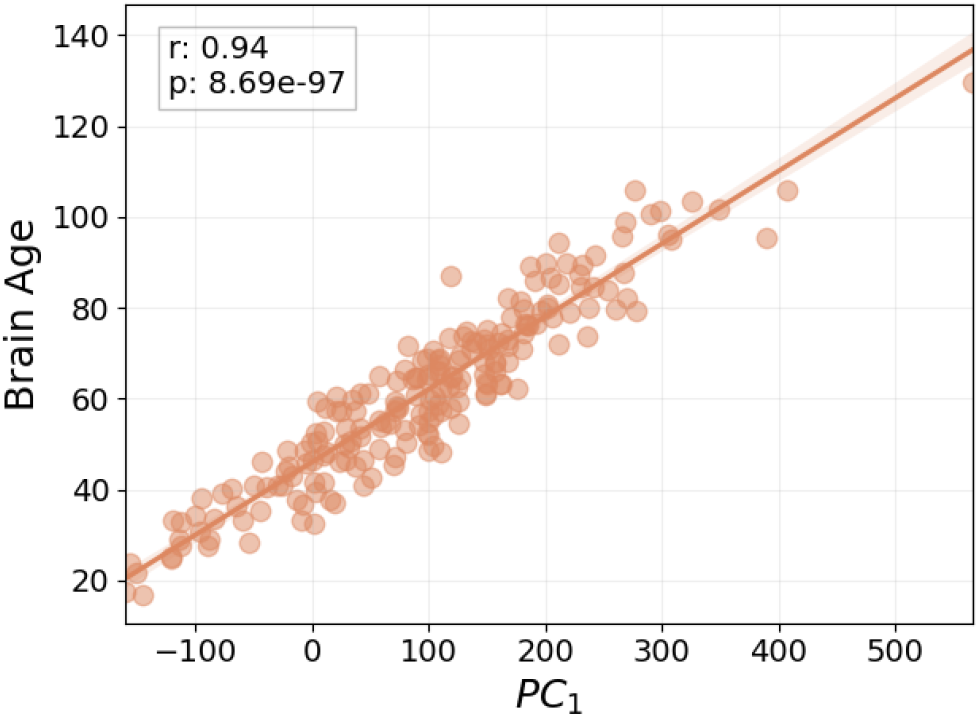
Scatterplot between the first principal component (*PC*_1_) and brain age in the MS_test dataset. The textbox describes the Pearson r statistic, along with the p-value.

## Discussion

This study aimed to investigate the potential of brain age, an intuitive metric of brain health, as a biomarker for information processing speed in Multiple Sclerosis. Our results suggest that brain age could be a promising candidate; it was substantially related to information processing speed, independent of chronological age. Moreover, we showed that brain age explained the majority of variance in brain volumetry by establishing a strong relationship with the first principal component of our total set of features.

In the past few years, most brain age-related research in MS was dedicated to establish clinical correlates of the Brain-Predicted Age Difference (BPAD)(11–13). Although BPAD might be regarded as a simplification of brain age, we in fact lose valuable information by subtracting two variables. Our results supports this statement in two ways. First, BPAD showed a weaker correlation with IPS than brain age. Second, brain age and chronological age both contributed unique information in explaining IPS in MS. Hence, in terms of clinical significance, BPAD remains at the level of proof-of-concept.

Although brain age is a fair choice for decoding processing speed in MS, we note that its performance was comparable to whole brain volume, as shown by Golan et al. 2020, reporting a correlation of r = 0.40 between whole brain volume and information processing speed(20). Nonetheless, brain age has an important advantage in contrast to any biological correlate of cognition in MS; it is easy to grasp as “how old a brain looks”, which facilitates communication with persons with MS. Constructing an uncomplicated message with minimal jargon contributes to optimally transferring medical information to patients, in turn optimising patient care(14).

However, an important hurdle in the path of brain age models to clinical practice is nicely illustrated by a statement in Ribeiro et al 2016: “if the users do not trust a model or a prediction, they will not use it”(24). First, to maximise trust of the MS clinician in our model, we used a model that is based on simple linear regression. The MS clinician has been familiarised with this method by decades of research adopting it for various purposes, e.g. studying the relation between MRI and cognitive performance in multiple sclerosis(25, 26). This contrasts with other studies on brain age in MS, mostly adopting models that are common in machine learning, but not in clinical research, such as Gaussian processes regression(13) and extreme gradient boosting(12). Second, trust in the prediction of our model, i.e. predicted brain age, was enhanced by showing that brain age explains the majority of the variance in MRI-derived volumetric features and sex, used to train our brain age model. This observation is logical in light of what is known about the aging brain, shrinking as people get older(27).

Our results imply that brain age has the potential to explain cognitive status in people with Multiple Sclerosis. Yet, the cross-sectional nature of the data limited us to investigate the potential of brain age to predict future decline in processing speed. Another limitation concerns the difference in segmentation method between the HC_train data and all test data. In HC_train, segmentation relied solely on T1-weighted images, whereas lesion filling in the T1-weighted image occurred in the test datasets with the use of FLAIR images. However, lesion filling had little impact on brain age predictions in the MS sample of Cole et al. 2020.(13) To ascertain that our results were not driven by this methodological issue, we recalculated brain age in the MS_test dataset on the same set of features, except that lesion volume was subtracted from white matter and whole brain volume. The results of this analysis, which can be consulted in the supplementary material (figure S3 and S4), indicate that brain age predictions were hardly affected by flair hyperintensity volume, and therefore do not explain the observations in this paper. Finally, our brain age model implemented a correction step to correct for the inherent over- and underestimation of younger and older subjects respectively. Since there appears to be no firm consensus about implementing this step, we reran the entire brain age pipeline without brain age correction, which did not alter the interpretation of this paper.

In summary, the methodology of a linear brain age model can be interpreted by clinicians, and its prediction by patients. Together with its potential to explain performance in information processing speed, predicted brain age could be a valuable clinical tool to analyse and communicate results from brain imaging data in MS.

## Supporting information

coi_disclosure

STROBE_checklist

## Data Availability

Data that was used to train our model was derived from publicly available repositories, of which the details can be found in the manuscript. Availability of the other data is subject to discussion with the senior author and may be limited by the ethics approval.

## Acknowledgements

The authors wish to thank Ann Van Remoortel and Jeroen Gielen for contributing to the data acquisition in the BRUMEG study.

## Supplementary Material

### Data characteristics HC_train dataset

**Table S1.**
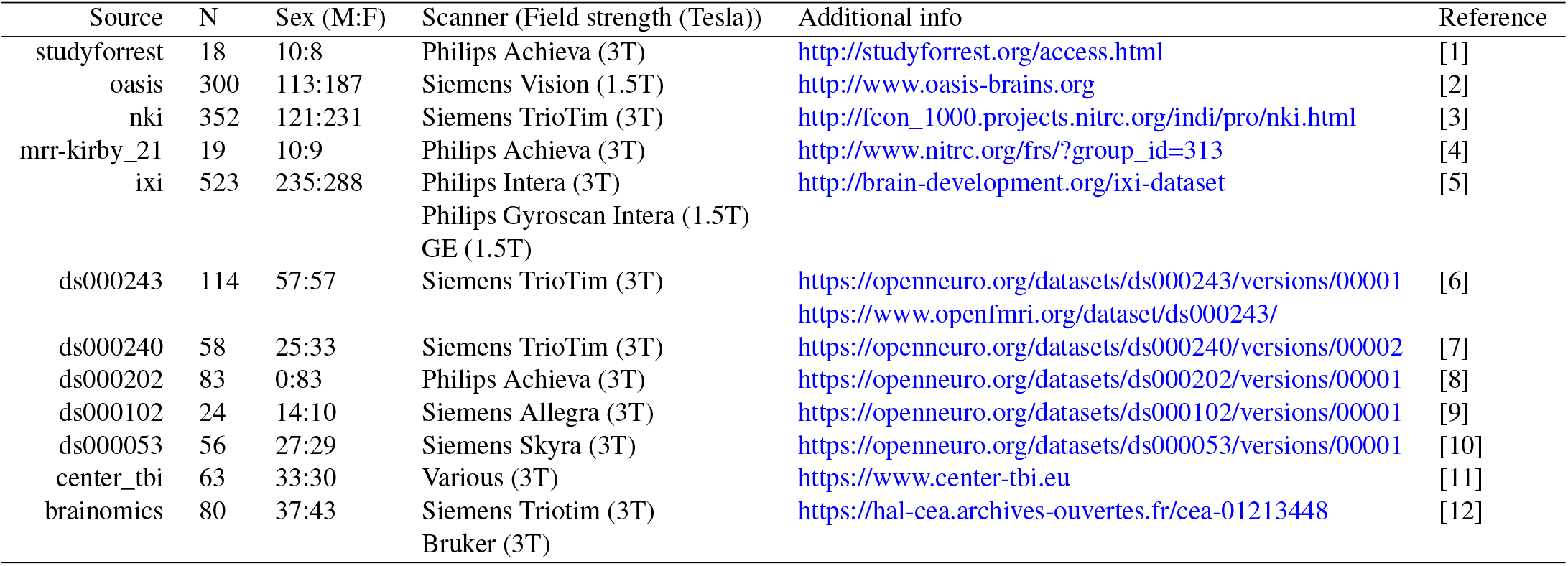
Characteristics of each data source in the HC_train dataset.

| Source | N | Sex (M:F) | Scanner (Field strength (Tesla)) | Additional info | Reference |
| --- | --- | --- | --- | --- | --- |
| studyforrest | 18 | 10:8 | Philips Achieva (3T) | http://studyforrest.org/access.html | [1] |
| oasis | 300 | 113:187 | Siemens Vision (1.5T) | http://www.oasis-brains.org | [2] |
| nki | 352 | 121:231 | Siemens TrioTim (3T) | http://fcon_1000.projects.nitrc.org/indi/pro/nki.html | [3] |
| mrr-kirby_21 | 19 | 10:9 | Philips Achieva (3T) | http://www.nitrc.org/frs/?group_id=313 | [4] |
| ixi | 523 | 235:288 | Philips Intera (3T)<br>Philips Gyroscan Intera (1.5T)<br>GE (1.5T) | http://brain-development.org/ixi-dataset | [5] |
| ds000243 | 114 | 57:57 | Siemens TrioTim (3T) | https://openneuro.org/datasets/ds000243/versions/00001<br>https://www.openfmri.org/dataset/ds000243/ | [6] |
| ds000240 | 58 | 25:33 | Siemens TrioTim (3T) | https://openneuro.org/datasets/ds000240/versions/00002 | [7] |
| ds000202 | 83 | 0:83 | Philips Achieva (3T) | https://openneuro.org/datasets/ds000202/versions/00001 | [8] |
| ds000102 | 24 | 14:10 | Siemens Allegra (3T) | https://openneuro.org/datasets/ds000102/versions/00001 | [9] |
| ds000053 | 56 | 27:29 | Siemens Skyra (3T) | https://openneuro.org/datasets/ds000053/versions/00001 | [10] |
| center_tbi | 63 | 33:30 | Various (3T) | https://www.center-tbi.eu | [11] |
| brainomics | 80 | 37:43 | Siemens Triotim (3T)<br>Bruker (3T) | https://hal-cea.archives-ouvertes.fr/cea-01213448 | [12] |

**Fig. S1.**
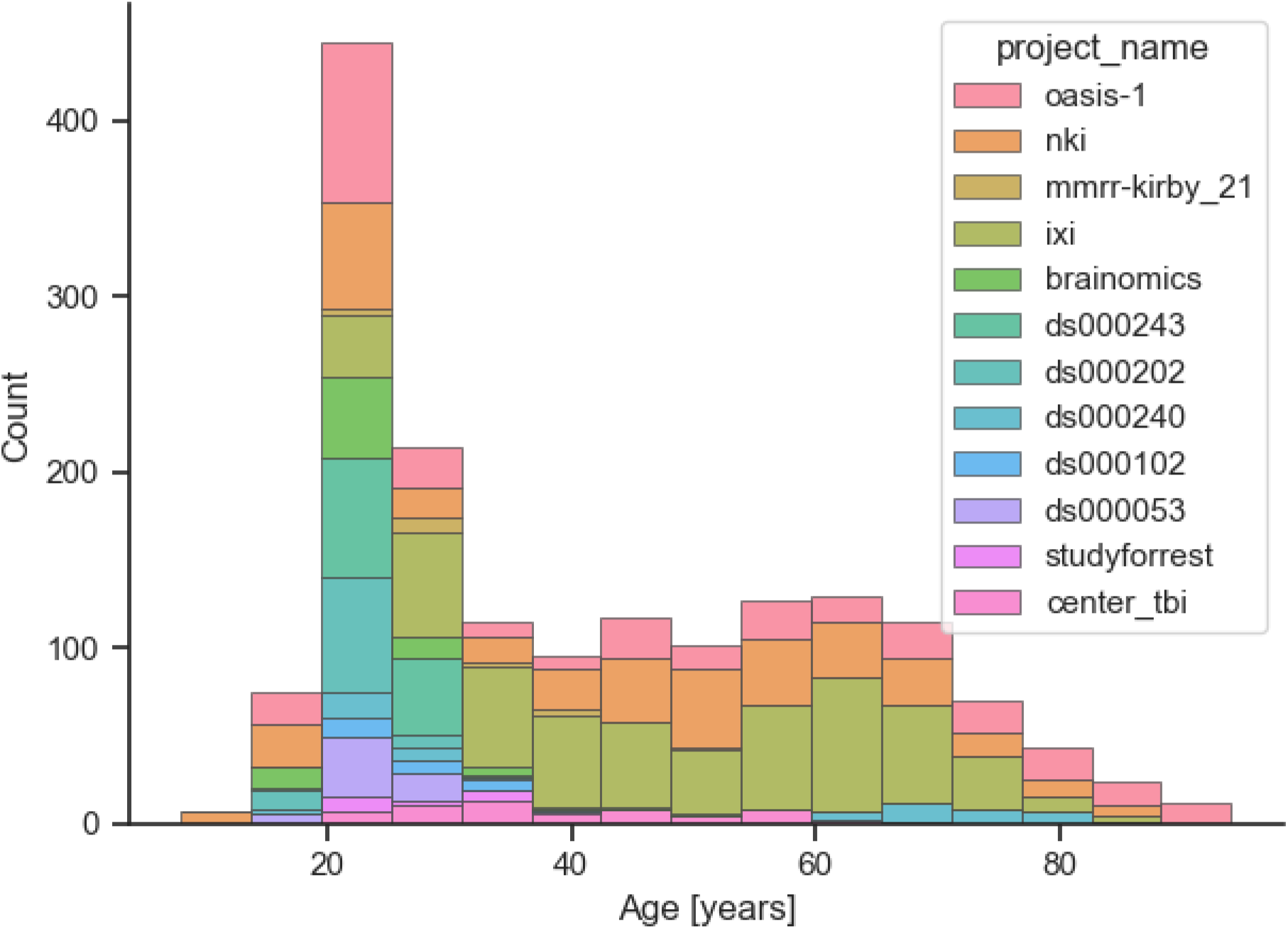
Histogram of the chronological ages per data source in the HC_train dataset

### Brain age correction

**Fig. S2.**
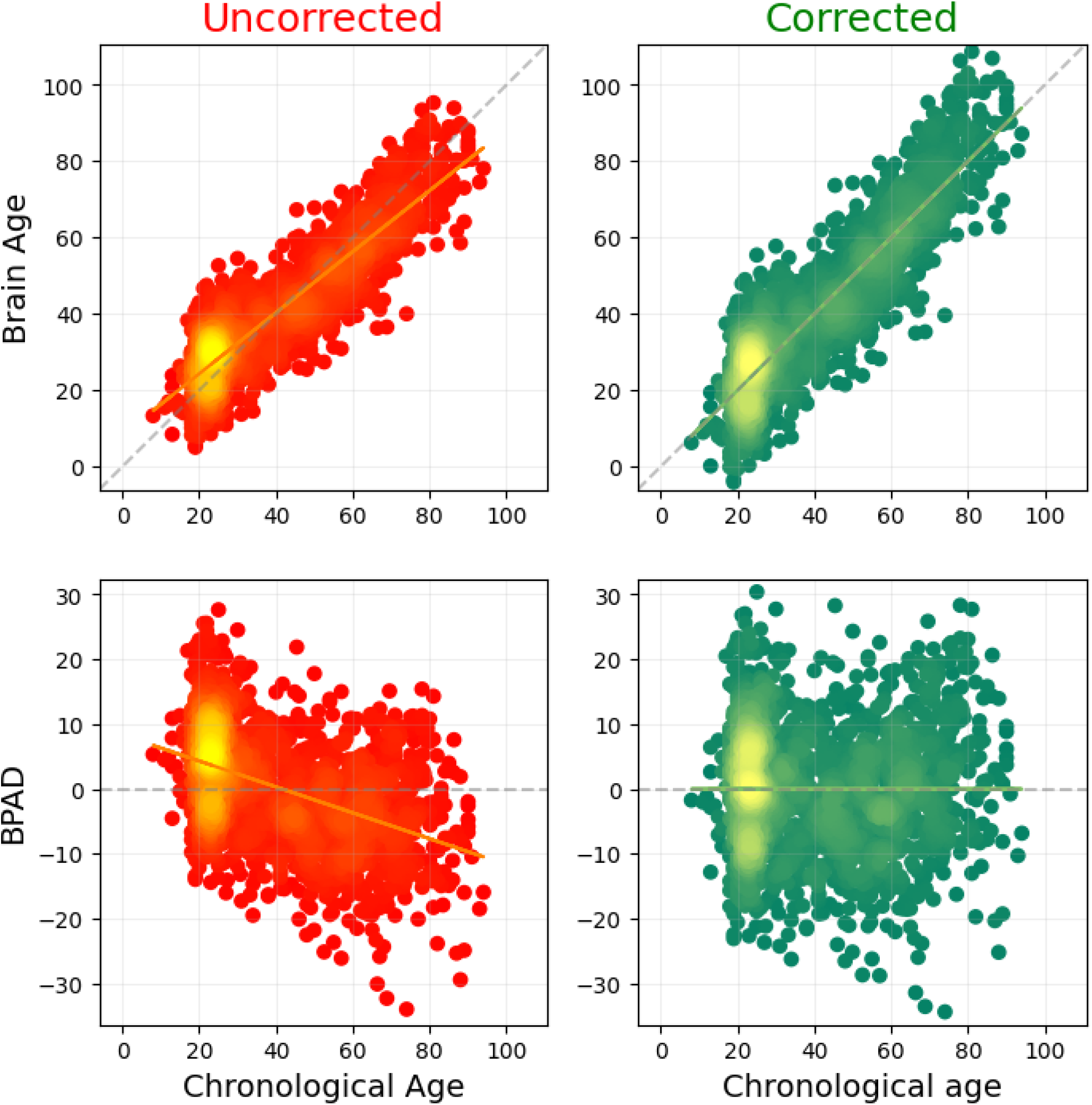
Brain age and BPAD as calculated with 10-fold cross-validation on the HC_train data. Uncorrected (raw) brain age and BPAD are indicated in red (regression line is indicated in orange), whereas corrected values are shown in green (regression line is indicated in light green). Both colormaps indicate higher density when yellower. Dotted lines were added to each plot as visual reference. For the upper plots, this is the line where Brain Age = Chronological Age, whereas for the lower plots, this is the line where BPAD = 0. Before correction, the regression lines do not coincide with the dotted lines, resulting in an overestimation of brain age in young individuals and an underestimation in older subjects. This is no longer the case after correction.

### Impact of lesions on brain age estimation

*BrainAge*_*lc*_ (lesion correction) and *BrainAge*_*n*−*lc*_ (no lesion correction) refer to the brain age that was calculated respectively with and without subtracting lesion volume from whole brain and white matter volume.

**Fig. S3.**
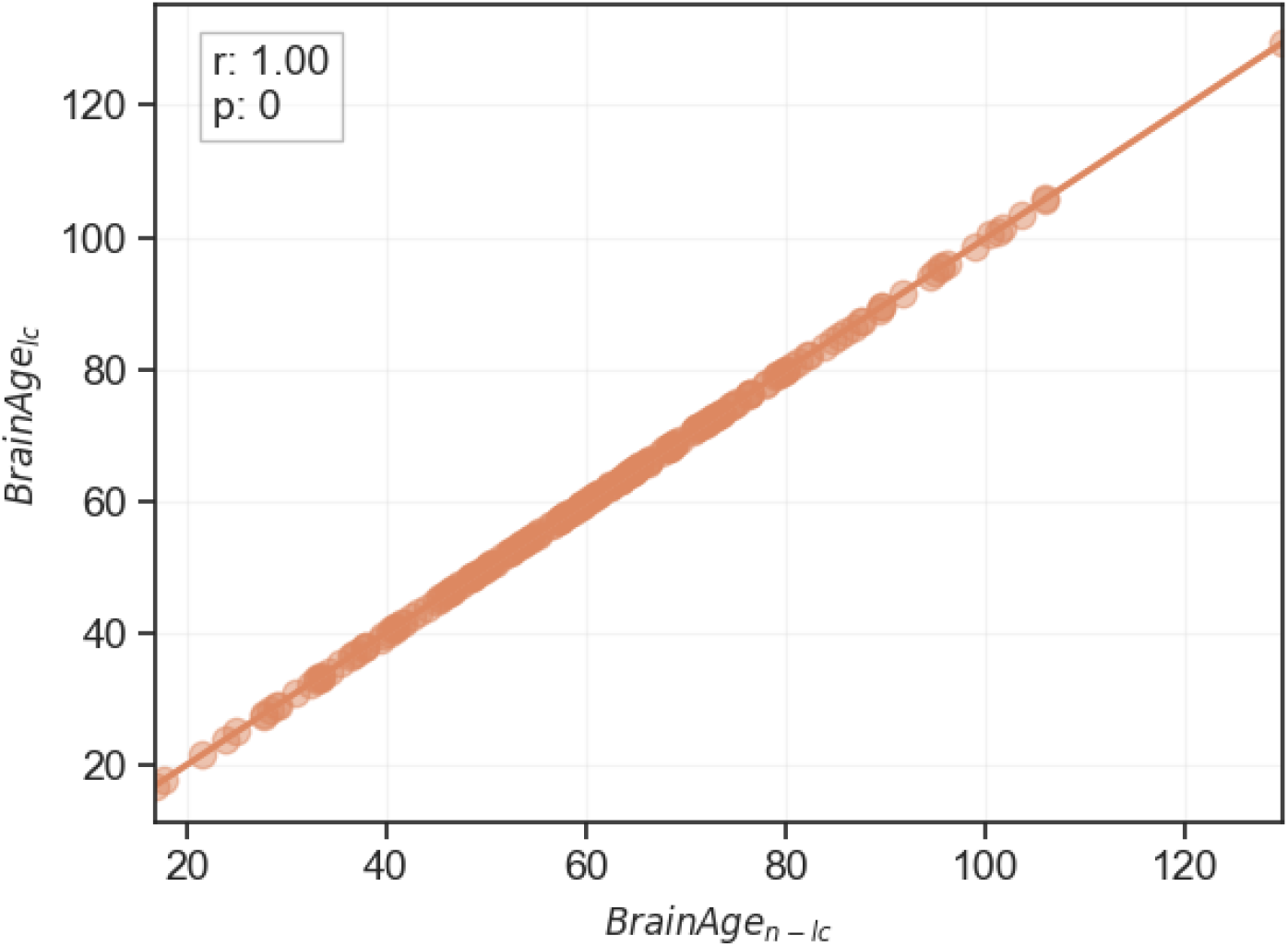
Scatterplot showing the relationship between *BrainAge*_*lc*_ and *BrainAge*_*n*−*lc*_.

**Fig. S4.**
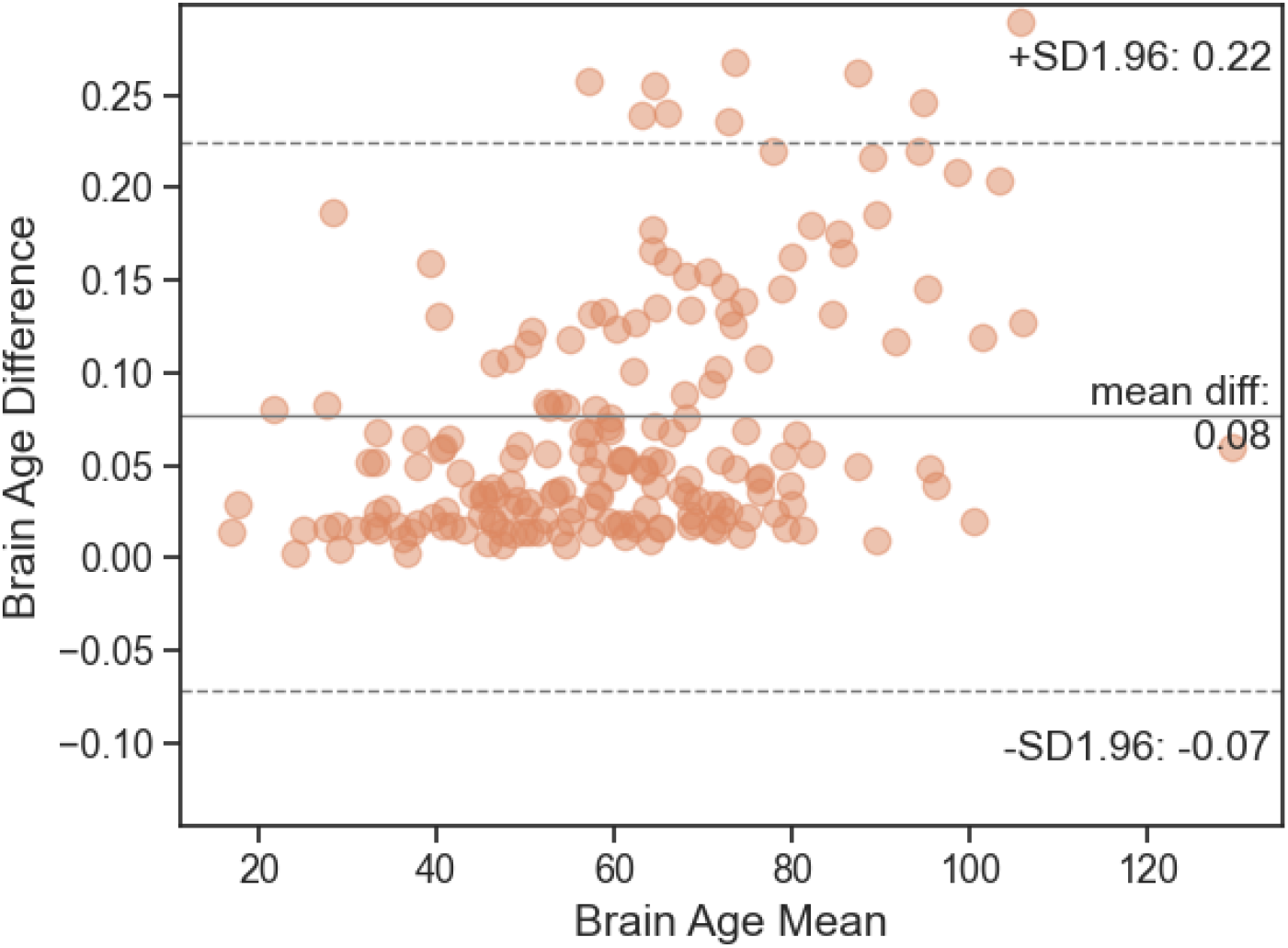
Bland-Altman plot showing the brain age difference (*BrainAge*_*n*−*lc*_ − *BrainAge*_*lc*_) for every brain age mean, calculated by (*BrainAge*_*n*−*lc*_ + *BrainAge*_*lc*_)*/*2. Both the mean and difference are expressed in terms of years.

## Preprint template acknowledgement

This preprint was created using a [LATEX template](https://www.overleaf.com/latex/templates/henriqueslab-biorxiv-template/nyprsybwffws) by Ricardo Henriques in the online LATEX editor [Overleaf](https://www.overleaf.com/). The authors wish to express their gratitude to Ricardo Henriques for publicly sharing this preprint template.

